# *In vivo* assessment of astrocyte reactivity in patients with progressive supranuclear palsy

**DOI:** 10.1101/2023.11.26.23298801

**Authors:** Kosei Hirata, Kiwamu Matsuoka, Kenji Tagai, Hironobu Endo, Harutsugu Tatebe, Maiko Ono, Naomi Kokubo, Yuko Kataoka, Asaka Oyama, Hitoshi Shinotoh, Keisuke Takahata, Takayuki Obata, Masoumeh Dehghani, Jamie Near, Kazunori Kawamura, Ming-Rong Zhang, Hitoshi Shimada, Hiroshi Shimizu, Akiyoshi Kakita, Takanori Yokota, Takahiko Tokuda, Makoto Higuchi, Yuhei Takado

## Abstract

Emerging evidence highlights the association of reactive astrocytes with neurodegenerative diseases. Although astrocytic pathology is a pathological hallmark of progressive supranuclear palsy (PSP), its role in the pathophysiology of the disease is not fully understood. In this study, we aimed to assess astrocyte reactivity *in vivo* in patients with PSP using magnetic resonance spectroscopy and plasma biomarkers. Furthermore, given the central role of astrocytes in brain energy metabolism and their glycolytic profile, which implies a preference for lactate production, we investigated alterations in brain energy metabolism by measuring brain lactate levels and examined their relationship with astrocyte reactivity.

We included 30 patients with PSP-Richardson’s syndrome and 30 healthy controls; in patients, tau deposition was confirmed via ^18^F-florzolotau-PET. Myo-inositol, an astroglial marker, and lactate were quantified in the anterior cingulate cortex (ACC) via magnetic resonance spectroscopy. The ACC was studied because previous functional imaging studies revealed its involvement as a distinctive feature of PSP. It involves in cognitive processes, such as executive function, which often exhibit deficits in patients with PSP. We measured plasma concentrations of glial fibrillary acidic protein (GFAP) as another astrocytic marker, neurofilament light chain (NfL), and tau phosphorylated at threonine 181. Reactive astrocytes, tau deposition, and synaptic loss in the ACC were assessed in post-mortem brain samples from another three patients with PSP with comparable disease durations to those of participants.

The level of myo-inositol in the ACC and the plasma GFAP were significantly higher in patients than in healthy controls; these increases were significantly associated with Frontal Assessment Battery and Mini-Mental State Examination scores, respectively. The lactate level in the ACC was high in patients and correlated significantly with high myo-inositol levels. The plasma NfL outperformed other biomarkers in discriminating patients from controls (area under the curve [AUC] = 0.95), followed by lactate and myo-inositol (AUC = 0.88 and 0.78, respectively). Histological analysis of the ACC in patients revealed evident reactive astrocytes despite mild tau deposition and no marked synaptic loss.

We found high levels of astrocyte biomarkers (myo-inositol in the ACC and plasma GFAP) in patients with PSP, suggesting astrocyte reactivity, and these biomarkers correlated with cognitive decline. Elevated myo-inositol levels were associated with high lactate levels, suggesting a link between reactive astrocytes and brain energy metabolism changes. Our results indicate that astrocyte reactivity in the ACC precedes pronounced tau pathology and neurodegenerative processes in the region and affects brain function in PSP.

## Introduction

Astrocytes are a type of glial cell that play a critical role in supporting and maintaining the function of the central nervous system.^1^ They undergo morphological and biochemical changes in response to injury, disease, and other insults, influencing the underlying pathophysiology.^2^ Recent studies have shed light on the role of such reactive astrocytes in the development and progression of neurodegenerative diseases.^3,4^ Progressive supranuclear palsy (PSP) is a neurodegenerative disorder characterised by the accumulation of four-repeat tau, leading to motor and cognitive impairments.^5^ In this disease, astrocytes exhibit tau-positive morphological changes and are known as “tufted astrocytes”.^5,6^ Although astrocytic tau pathology is a pathological hallmark of the disease, and reactive astrocytes have been noted in histopathological studies,^6^ the role and importance of astrocytes in the pathophysiology of PSP remains unclear.

*In vivo* assessment of astrocyte reactivity would provide valuable insight into the pathophysiology of PSP. Glial fibrillary acidic protein (GFAP) is predominantly expressed in astrocytes, and its elevated expression has been considered a marker for astrocyte reactivity.^2,4^ Plasma GFAP is an emerging biomarker for Alzheimer’s disease (AD), associated with cognitive decline and neurodegeneration.^7^ Several studies have revealed significantly elevated concentrations of plasma GFAP in patients with PSP compared to those in healthy controls (HCs).^8,9^ Nevertheless, peripheral biomarkers cannot be used to determine alterations within specific brain regions. Proton magnetic resonance spectroscopy (MRS) enables the non-invasive quantification of tissue metabolites, providing insights into distinct cellular and molecular alterations within specific brain areas. The use of myo-inositol, a metabolite detectable by MRS, holds promise for the assessment of astrocyte reactivity.^10,11^ This metabolite, which mainly found in glial cells, is reportedly elevated in reactive astrogliosis.^12–14^ In patients with AD, increased myo-inositol levels in the posterior cingulate cortex (PCC) region, a hub of the default mode network (DMN), are reportedly noted from the early stages of the disease.^15^ Few MRS studies on the neurometabolic alterations in patients with PSP have been published.^16,17^ A recent study in which whole-brain MRS was used revealed increased levels of myo-inositol in the frontal cortex of patients with PSP compared to those in the frontal cortex of HCs, although its clinical and pathophysiological relevance were not addressed.^18^ In patients with PSP, compared with HCs, decreased glucose utilization in the frontal lobe, including the anterior cingulate cortex (ACC), midbrain, basal ganglia, and thalamus, has been demonstrated in PET studies in which ^18^F-fluorodeoxyglucose (FDG) was used.^19,20^ Additionally, a brain-perfusion single-photon emission CT (SPECT) study revealed hypoperfusion of the ACC and medial frontal cortex in PSP.^21^ These results suggest that ACC involvement is a distinct characteristic of PSP. The ACC is another pivotal region of the DMN, playing an important role in cognitive processes such as executive function,^22^ which are often impaired in patients with PSP.^23^ Therefore, in this study, we aimed to examine the myo-inositol levels in the ACC and its association with cognitive function scores in patients with PSP.

Astrocytes are central regulators of energy metabolism in the brain.^24^ Considering their role in energy homeostasis, astrocyte reactivity may lead to changes in brain energy metabolism. A recent PET study, which combined acetate- and FDG-PET, demonstrated that reactive astrogliosis is associated with decreased glucose utilization in AD.^25^ With regard to brain energy metabolism related to astrocytes, lactate has emerged as a key molecule, given the reliance of astrocytes on glycolysis and their propensity to produce lactate.^24^ We recently demonstrated increased lactate levels in the PCC and their association with increased myo-inositol levels by using an advanced MRS sequence in patients with AD.^11^ This study used the same MRS technique to quantify both lactate and myo-inositol levels in the ACC to investigate the potential impact of reactive astrocytes on changes in brain energy metabolism in PSP.

In light of recent research highlighting the role of astrocytes in neurodegenerative diseases and the characteristic astrocytic pathology of PSP, we hypothesised that astrocyte dysfunction contributes to the pathophysiology of PSP. We tested this hypothesis in the current study by assessing astrocyte reactivity *in vivo* and its relationship with cognitive function scores, brain energy metabolism, and tau pathology by using MRS, plasma biomarkers, and tau-PET, which were verified via histopathological analysis.

## Materials and methods

### Participants

Participants with probable PSP with Richardson’s syndrome, diagnosed based on the International Parkinson and Movement Disorder Society clinical diagnostic criteria for PSP,^26^ were prospectively recruited at the affiliated hospitals of the National Institute of Radiological Sciences (NIRS) and National Institutes for Quantum Science and Technology (QST) from February 2018 to February 2022. At the initial screening, all participants with PSP were verified as tau-positive by visual evaluation of ^18^F-florzolotau-PET images, confirming the PSP diagnosis with high certainty.^27^ Age- and sex-matched HCs were recruited from the volunteer association of the NIRS-QST, all of whom did not have a history of neurological or psychiatric disorders. The cognitive function of all participants was evaluated using the Mini-Mental State Examination (MMSE) for global cognition and the frontal assessment battery (FAB) for frontal lobe dysfunction. Finally, a total of 30 patients with PSP and 30 HC individuals were enrolled in the study.

This study was approved by the Institutional Review Board of QST. Written informed consent was obtained from all participants and/or from the spouses or other close family members of participants who were cognitively impaired.

### MRI/MRS data acquisition

MRI and MRS were performed using a 3.0-T scanner (MAGNETOM Verio; Siemens Healthcare) with a 32-channel receiving head coil. We obtained structural, three-dimensional, T1-weighted images by using the magnetization-prepared, rapid gradient-echo sequence with the following parameters: echo time (TE) = 1.95 ms, repetition time (TR) = 2,300 ms, inversion time = 900 ms, field of view = 250 mm, flip angle = 9°, acquisition matrix = 512 × 512, and an axial slice thickness of 1 mm. Single-voxel MRS data were collected using a short-TE spin-echo full-intensity acquired localised single-voxel spectroscopy (SPECIAL) sequence with TE of 8.5 ms, TR of 3,000 ms, and 128 averages, which was designed to capture a full-intensity signal in a designated volume of interest (VOI) by using an ultra-short TE.^28^ The ACC was the chosen location for the VOI (30 × 20 × 20 mm^3^) (Fig. 1A).

**Figure 1.**
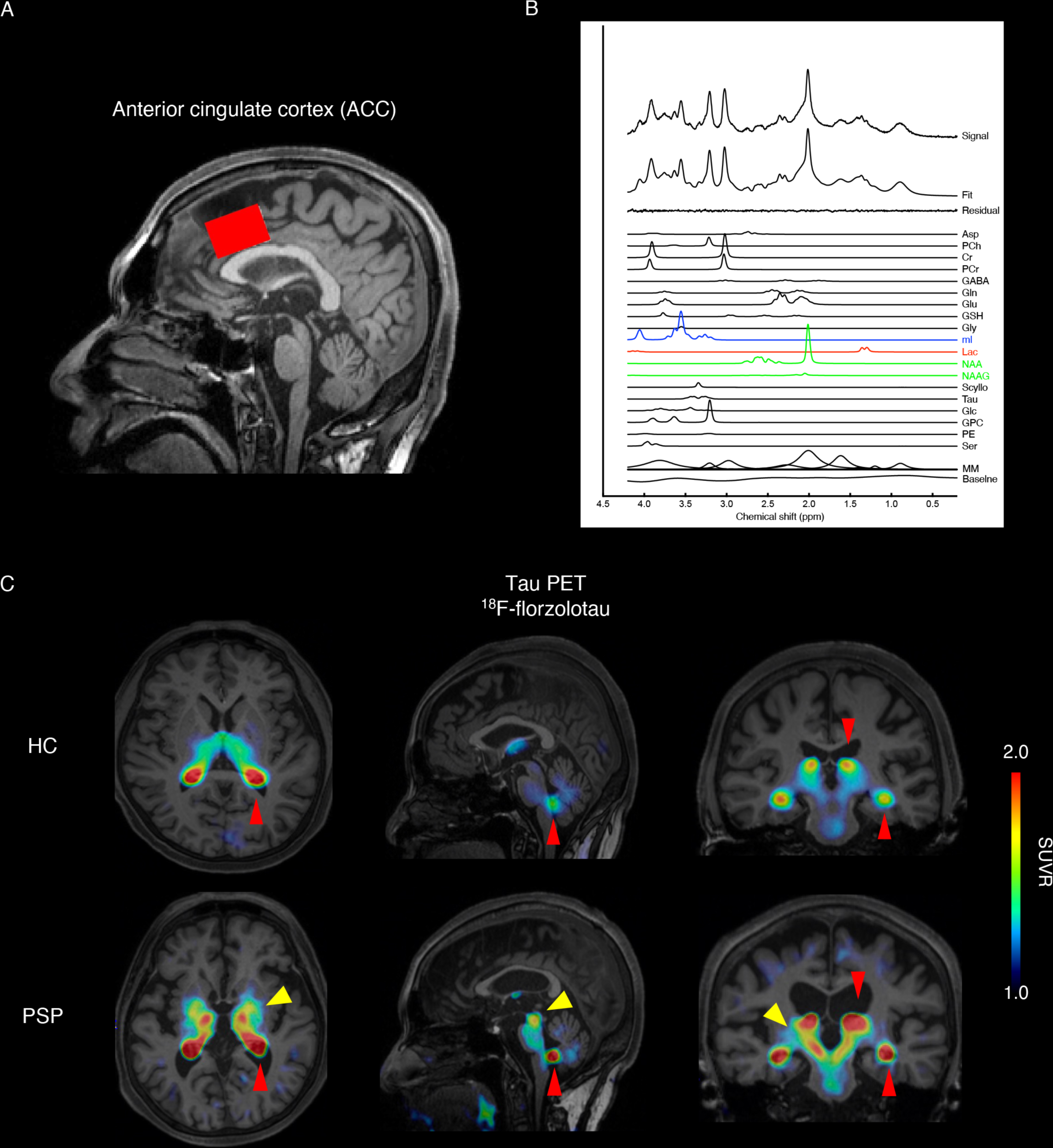
MRS voxel location, example spectra, and representative tau-PET images using ^18^F-florzolotau. (**A**) A representative single voxel, sized 30 × 20 × 20 mm^3^, is positioned in the anterior cingulate cortex and superimposed on a T1-weighted image. (**B**) An example MRS spectrum derived from the voxel of a patient with PSP. The MRS data, obtained using the SPECIAL sequence, were analysed with the LCModel. The LCModel spectral fit, fit residual, macromolecules, baseline, and individual metabolite fits for myo-inositol (in blue), lactate (in red), and tNAA (in green) are displayed. (**C**) Representative ^18^F-florzolotau tau-PET images of a HC participant and one from the PSP group. In PSP, enhanced ^18^F-florzolotau binding was observed in the subthalamic nucleus, thalamus, basal ganglia, and midbrain, as indicated by the yellow arrowheads. Red arrowheads point to non-specific binding in the choroid plexus. HC = healthy control; MM = macromolecules; MRS = magnetic resonance spectroscopy; PSP = progressive supranuclear palsy; SPECIAL sequence = short-TE spin-echo full-intensity acquired localised single-voxel spectroscopy sequence; SUVR = standardised uptake value ratio; tNAA = total N-acetyl-aspartate.

### MRS analysis

A weighted combination of receiver channels was performed using the FID-A toolkit, followed by removal of motion-corrupted averages, spectral registration for frequency and phase drift correction, and alignment of subspectra prior to subtraction.^29^ MRS data were processed using LCModel software (Stephen Provencher, Inc.) for a linear combination of model spectra with simulated basis functions.^30^ We used a neurochemical basis set that incorporated nine macromolecular basis functions, each sourced from the individual peaks in a macromolecular spectrum obtained by the summation of experimentally acquired metabolite-nulled spectra from six healthy adult volunteers. A representative spectrum is illustrated in Fig. 1B. The average ± standard deviation (SD) spectral signal-to-noise ratio (SNR) and line width (LCModel output) were 78.4 ± 16.9 and 3.91 ± 1.00 Hz, respectively. All MRS data met the SNR and line-width quality criteria.^31^ In this study, we evaluated the levels of myo-inositol, lactate, and total N-acetyl-aspartate (tNAA: NAA + N-acetyl-aspartyl-glutamate [NAAG]) as neuronal biomarkers. Owing to severe artifact interference leading to data absence, lactate evaluation was not performed in one individual in the HC group. The Cramér-Rao lower bound (CRLB) provided by LCModel was used to assess the reliability of neurochemical quantification for each metabolite. A cut-off of 30% CRLB averaged over all scans was applied in this study. Excluding individual data with elevated CRLB values, especially for low-concentration metabolites, is not recommended, as it may bias the cohort mean concentrations.^32^ Tissue water served as the internal reference for concentration quantification. Partial volume effects were corrected by extracting the grey matter (GM), white matter (WM) and CSF fractions within the VOIs derived from the segmentation of T1-weighted images by using Gannet 3.0 software.^33^ The estimated water concentrations for GM, WM, and CSF were 35,880, 43,300, and 55,556 mM, respectively. Water concentrations were subsequently corrected on the basis of the partial volume fractions of GM, WM, and CSF by using an equation described previously.^34^ Metabolite concentrations (μmol/g) were divided by the GM and WM fractions to account for CSF in the VOI.

### Voxel-based morphometry

Data pre-processing was performed using Statistical Parametric Mapping software (SPM12, Wellcome Department of Cognitive Neurology). T1-weighted images were spatially normalised to the Montreal Neurological Institute space, specifically the East Asian brain T1-weighted images from the International Consortium for Brain Mapping, by using the Diffeomorphic Anatomical Registration Through Exponentiated Lie Algebra algorithm with an 8-mm full width at half maximum isotropic Gaussian kernel. Voxel-based morphometric (VBM) analysis was conducted on the combined, pre-processed GM and WM segments of the T1-weighted images derived from the SPM segmentation process. The total brain volume served as a confounding covariate in an analysis of covariance. A statistical significance threshold of *P* < 0.001 was applied to identify clusters exceeding the expected number of voxels per cluster.

### PET acquisition and analysis

Cerebral Aβ and tau depositions were assessed using ^11^C-Pittsburgh Compound-B (PiB) and ^18^F-florzolotau-PET, respectively. We recently developed the PET radiotracer, ^18^F-florzolotau, to visualise tau lesions across various tauopathies, encompassing the detection of mixed three-repeat and four-repeat tau fibrils in AD and four-repeat tau aggregates in PSP and corticobasal degeneration.^27^ ^11^C-PiB and ^18^F-florzolotau were radiosynthesised according to the protocols in our previous studies.^27^ The participants received an intravenous injection of approximately 185 MBq of ^18^F-florzolotau and were scanned for 90–110 min after injection. All images were collected using the Biograph mCT Flow system (Siemens Healthcare). ^11^C-PiB (injected dose: approximately 555 MBq) PET was performed, scanning participants for 50–70 min after injection by using the ECAT EXACT HR+ Scanner (CTI PET Systems, Inc.), Biograph mCT Flow system, or Discovery MI (GE Healthcare). The PET images were reconstructed using a filtered back projection algorithm. PET images were analysed using PMOD software version 3.8 (PMOD Technologies Ltd.). Images were motion-corrected and co-registered to the corresponding T1-weighted images of the participants. Standardised uptake value ratio images were generated using the cerebellar GM as the reference region. For the reference region, we executed surface-based cortical reconstruction and subcortical volumetric segmentation of T1-weighted images via FreeSurfer software (version 6.0.0; http://surfer.nmr.harvard.edu), as detailed in our previous research.^35^ Representative PET images of ^11^C-PiB and ^18^F-florzolotau are presented in Fig. 1C. The median values for Aβ-PET and tau-PET retention in the ACC were calculated from the VOIs in the GM and WM corresponding to the MRS VOIs by using Gannet 3.0 software.

### Immunoassay protocols

Blood samples were evaluated for the plasma concentrations of GFAP, neurofilament light chain (NfL), and tau phosphorylated at threonine 181 (p-tau 181). Owing to inadequate quantities for assessment, two samples from the HC group and three from the PSP group were excluded from the analysis. The Simoa HD-X Analyzer was employed to analyse these samples by using commercially available kits from a single batch, as per the manufacturer’s guidelines (Quanterix).^36^ All samples were analysed in duplicate with the same lot of standards.

### Histological examination

We pathologically examined another three autopsy-confirmed cases of PSP-Richardson’s syndrome with disease duration (case 1, 61-65 years old; case 2, 81-85 years old; case 3, 76-80 years old) comparable to this study and three age-matched HCs from different studies. None of the patients with PSP or HCs showed pathological findings indicative of complications arising from AD.^37,38^ In PSP-2 and PSP-3, a few α-synuclein-positive Lewy bodies were observed in the brainstem, with a distribution pattern corresponding to the brainstem-predominant type of Lewy-related pathology.^39^ In PSP-2, a small number of argyrophilic grains were observed in the amygdaloid nucleus, with a distribution pattern corresponding to Stage I.^40^ There were no α-synuclein-positive Lewy bodies or argyrophilic grains in the ACC in patients with PSP or HCs. Accordingly, 4-μm-thick sections were cut from formalin-fixed, paraffin-embedded tissue blocks containing the ACC. Immunohistochemical analyses were performed as previously described,^41,42^ using mouse monoclonal antibodies against p-tau (AT8; Thermo Fisher Scientific, MN1020; 1:200) and synaptophysin (Leica Biosystems, NCL-SYNAP-299; 1:100), and using a rabbit polyclonal antibody against GFAP (Invitrogen, PA5-16291; 1:400).

### Statistical analysis

Intergroup differences were assessed using independent-samples t-tests for parametric data or Mann-Whitney tests for non-parametric data. Categorical data were analysed using chi-square tests. Partial correlation analysis, adjusted for age and sex, was conducted to evaluate correlations between clinical characteristics, metabolite levels, and plasma biomarkers. All statistical tests were two-tailed, and *P*-values of <0.05 denoted statistical significance; the Benjamini-Hochberg method was used for false discovery rate correction for multiple comparisons.^43^ The ability of each MRS and blood biomarker to discriminate between HC and PSP was determined using the area under the receiver operating characteristic curve (AUC) values. Statistical analyses were performed using IBM SPSS Statistics version 27 (IBM Corp.) and GraphPad Prism 9 software (GraphPad Software).

## Results

### Demographic and clinical data

The demographic and clinical characteristics of the participants are summarised in Table 1. The study included 30 HC individuals and 30 individuals diagnosed with probable PSP with Richardson’s syndrome. The groups did not differ in terms of age or sex. The average disease duration for the participants with PSP was 3.1 ± 1.9 years. The PSP group exhibited worse cognitive function, as demonstrated by lower total scores on the MMSE and FAB, than the HC group (*U* = 138, *P* < 0.001 and *U* = 59.5, *P* < 0.001, respectively). Among patients with PSP, ^18^F-florzolotau imaging revealed elevated radiotracer signals in the subthalamic nucleus, neighbouring thalamic and basal ganglia regions, and midbrain, consistent with the distribution patterns of tau pathology in PSP (Fig. 1C).^6^ All participants had negative results on amyloid-PET scans.

**Table 1.**
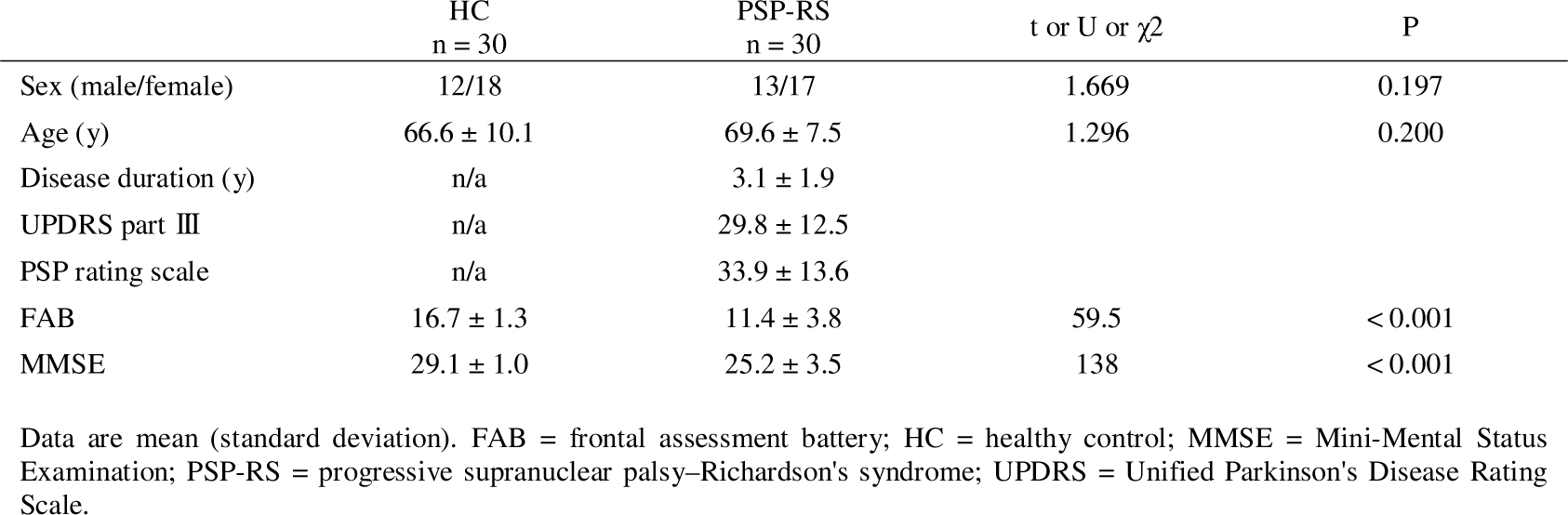
Baseline Demographics and Clinical Characteristics.

| | HC<br>n = 30 | PSP-RS<br>n = 30 | t or U or $\chi^2$ | P |
| --- | --- | --- | --- | --- |
| Sex (male/female) | 12/18 | 13/17 | 1.669 | 0.197 |
| Age (y) | 66.6 ± 10.1 | 69.6 ± 7.5 | 1.296 | 0.200 |
| Disease duration (y) | n/a | 3.1 ± 1.9 | | |
| UPDRS part III | n/a | 29.8 ± 12.5 | | |
| PSP rating scale | n/a | 33.9 ± 13.6 | | |
| FAB | 16.7 ± 1.3 | 11.4 ± 3.8 | 59.5 | < 0.001 |
| MMSE | 29.1 ± 1.0 | 25.2 ± 3.5 | 138 | < 0.001 |

Data are mean (standard deviation). FAB = frontal assessment battery; HC = healthy control; MMSE = Mini-Mental Status Examination; PSP-RS = progressive supranuclear palsy-Richardson's syndrome; UPDRS = Unified Parkinson's Disease Rating Scale.

### MRS results

The MRS metabolites assessed in the ACC included myo-inositol, lactate, and tNAA (Table 2 and Fig. 2A). The average ± SD CRLBs for myo-inositol, lactate, and tNAA were 3.7% ± 1.0%, 15.9% ± 6.5%, and 1.8% ± 0.7%, respectively, all satisfying the set criteria. Myo-inositol levels in the PSP group were significantly higher than in the HC group (*t* = 4.379, *P* < 0.001). The PSP group exhibited elevated lactate levels compared to the HC group (*U* = 106.5, *P* < 0.001). The tNAA levels did not significantly differ between the two groups (*U* = 330, *P* = 0.077). Subsequently, we investigated whether astrocytic alterations are linked to elevated lactate levels by assessing the association between myo-inositol and lactate levels. Higher levels of myo-inositol were associated with higher lactate levels (*r* = 0.603, *P* < 0.001 for all participants, *r* = 0.445, *P* = 0.018 for the PSP group) (Fig. 2B).

**Figure 2.**
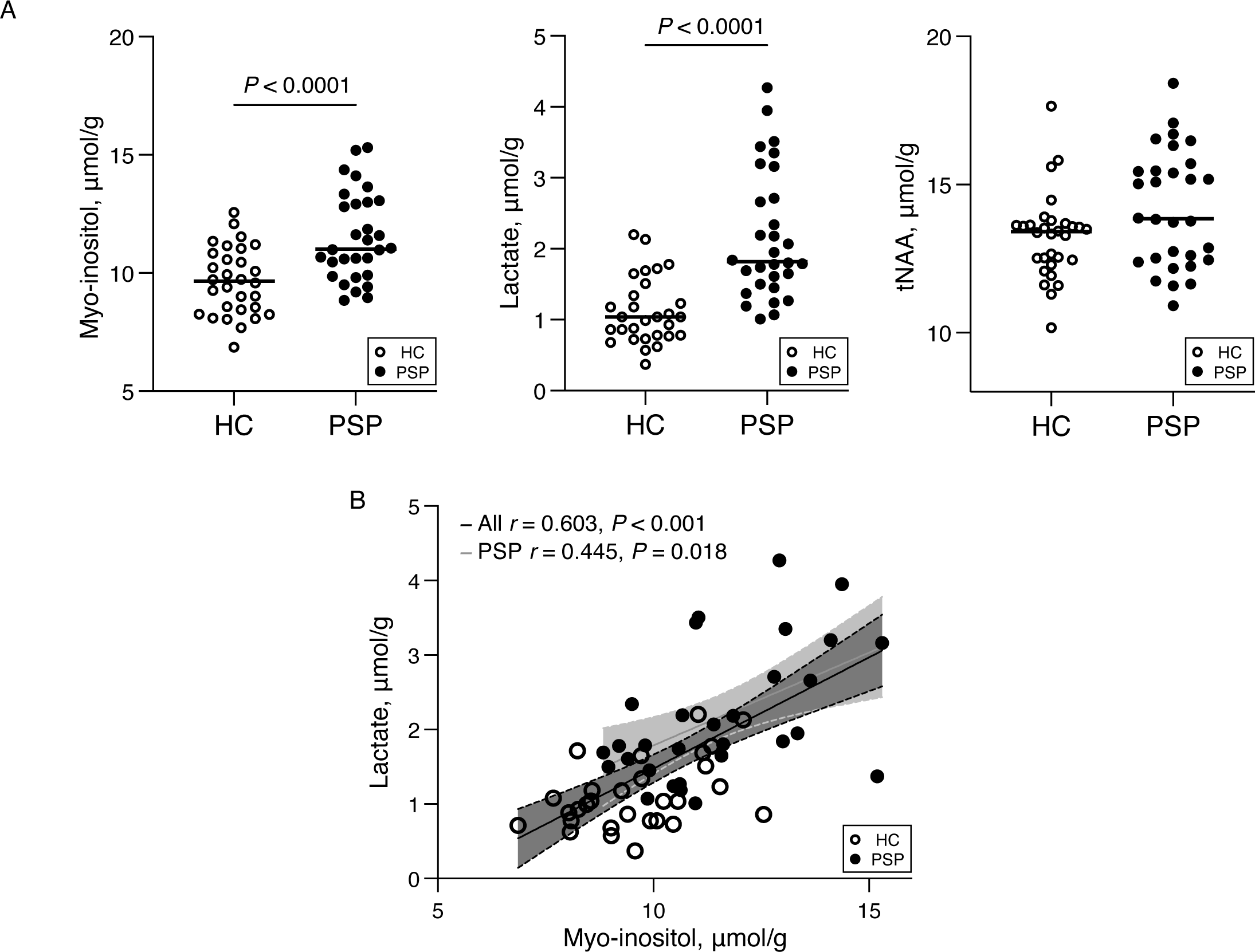
Scatter plots of the MRS metabolite levels for HCs and participants with progressive supranuclear palsy, and the association between myo-inositol and lactate levels. (**A**) Concentrations of myo-inositol, lactate, and tNAA in the anterior cingulate cortex for the HC and PSP groups. Bars represent median values. (**B**) The association between myo-inositol and lactate levels in the anterior cingulate cortex was assessed using a partial correlation analysis, adjusted for age and sex. Linear regression lines and a 95% confidence interval, were derived from the unadjusted analysis for all participants and specifically for participants with PSP, represented by black and grey lines, respectively. HC = healthy controls; MRS = magnetic resonance spectroscopy; PSP = progressive supranuclear palsy; tNAA = total N-acetyl-aspartate.

**Table 2.** Group Comparisons of MRS metabolites and plasma biomarkers.

|  | HC | PSP-RS | t or U | P |
| --- | --- | --- | --- | --- |
| **MRS metabolites ($\mu\text{mol/g}$)** |  |  |  |  |
| Myo-inositol | $9.7 \pm 1.4$ | $11.5 \pm 1.9$ | 4.379 | $< 0.001$ |
| Lactate | $1.1 \pm 0.5$ | $2.2 \pm 0.9$ | 106.5 | $< 0.001$ |
| tNAA | $13.2 \pm 1.4$ | $14.2 \pm 2.0$ | 330 | 0.077 |
| **Plasma biomarkers (pg/ml)** |  |  |  |  |
| GFAP | $105.5 \pm 41.9$ | $144.1 \pm 65.6$ | 246 | 0.039 |
| NfL | $18.0 \pm 8.2$ | $58.0 \pm 35.9$ | 37 | $< 0.001$ |
| p-tau 181 | $1.7 \pm 0.7$ | $2.1 \pm 0.9$ | 263 | 0.080 |

Data are mean (standard deviation). GFAP = glial fibrillary acidic protein; HC = healthy control; MRS = magnetic resonance spectroscopy; NfL = neurofilament light chain; PSP-RS = progressive supranuclear palsy–Richardson's syndrome; p-tau181 = tau phosphorylated at threonine 181; tNAA = total N-acetyl-aspartate.

### MRS and plasma biomarkers

Next, we examined the plasma biomarker changes between the two groups and associations between myo-inositol and plasma biomarkers. The plasma concentrations of GFAP and NfL were significantly higher in the PSP group than those in the HC group (*U* = 246, *P* = 0.039 for GFAP; *U* = 37, *P* < 0.001 for NfL) (Table 2 and Fig. 3A). The concentrations of plasma p-tau 181 did not differ between the HC and PSP groups (*U* = 263, *P* = 0.080). The myo-inositol level correlated significantly with the plasma NfL concentration but not with plasma GFAP (*r* = 0.290, *P* = 0.035 for plasma NfL; *r* = 0.249, *P* = 0.072 for plasma GFAP) (Fig. 3B). We further compared the diagnostic value of MRS and plasma biomarkers in identifying PSP. Plasma NfL yielded the highest accuracy in discriminating patients with PSP from HCs, with an AUC of 0.95 (Fig. 3C). The MRS biomarkers, myo-inositol and lactate yielded moderate accuracy in discriminating between the groups (AUC = 0.78 for myo-inositol; AUC = 0.88 for lactate), whereas plasma GFAP and p-tau 181 yielded AUCs of 0.67 and 0.64, respectively.

**Figure 3.**
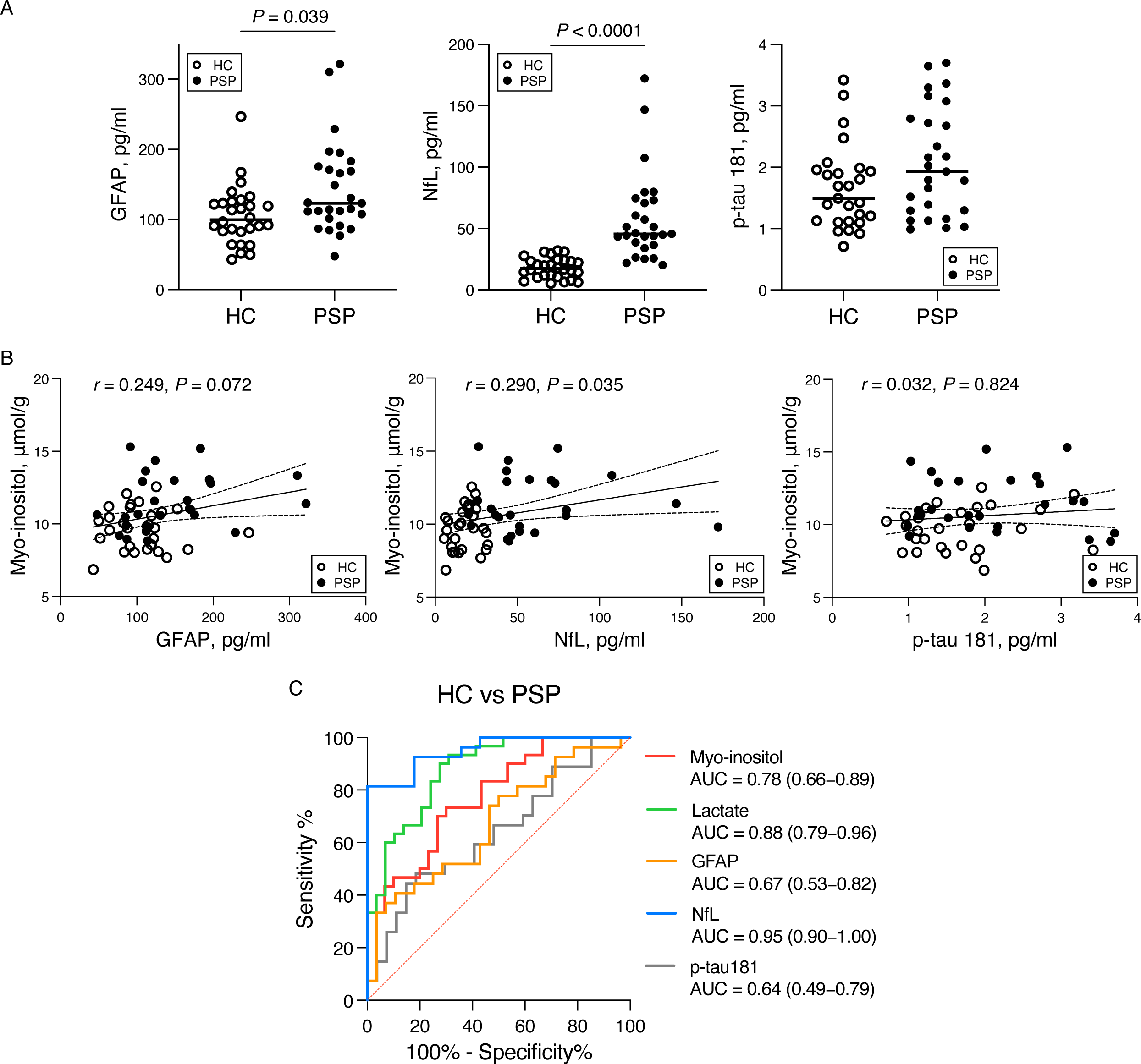
Scatter plots of plasma biomarker concentrations, associations between myo-inositol and plasma biomarkers, and ROC curves. (**A**) Concentrations of plasma GFAP and NfL in the HC and PSP groups. Bars indicate median values. (**B**) Associations between myo-inositol in the anterior cingulate cortex and plasma biomarkers were examined by partial correlation analysis adjusted for age and sex. A linear regression line with a 95% confidence interval was obtained from the unadjusted analysis. (**C**) ROC curves illustrating the ability of each MRS and blood biomarker to distinguish between HCs and patients with PSP, accompanied by the respective AUC values with their 95% confidence intervals. AUC = area under the ROC curve; HC = healthy control; MRS = magnetic resonance spectroscopy; GFAP = glial fibrillary acidic protein; NfL = neurofilament light chain; PSP = progressive supranuclear palsy; p-tau 181= tau phosphorylated at threonine 181; ROC; receiver operating characteristic.

### Associations of astrocyte biomarkers with cognitive decline in progressive supranuclear palsy

We assessed the clinical relevance of astrocyte reactivity in PSP by investigating the association between the astrocyte biomarkers examined in this study and cognitive function scores determined using the MMSE and FAB. Higher myo-inositol levels in the ACC were significantly correlated with lower FAB scores (*r* = −0.537, *P* = 0.003) (Fig. 4). An increased plasma GFAP concentration was significantly associated with a decreased MMSE score but not with the FAB score (*r* = −0.398, *P* = 0.049 for MMSE; *r* = −0.364, *P* = 0.074 for FAB).

**Figure 4.**
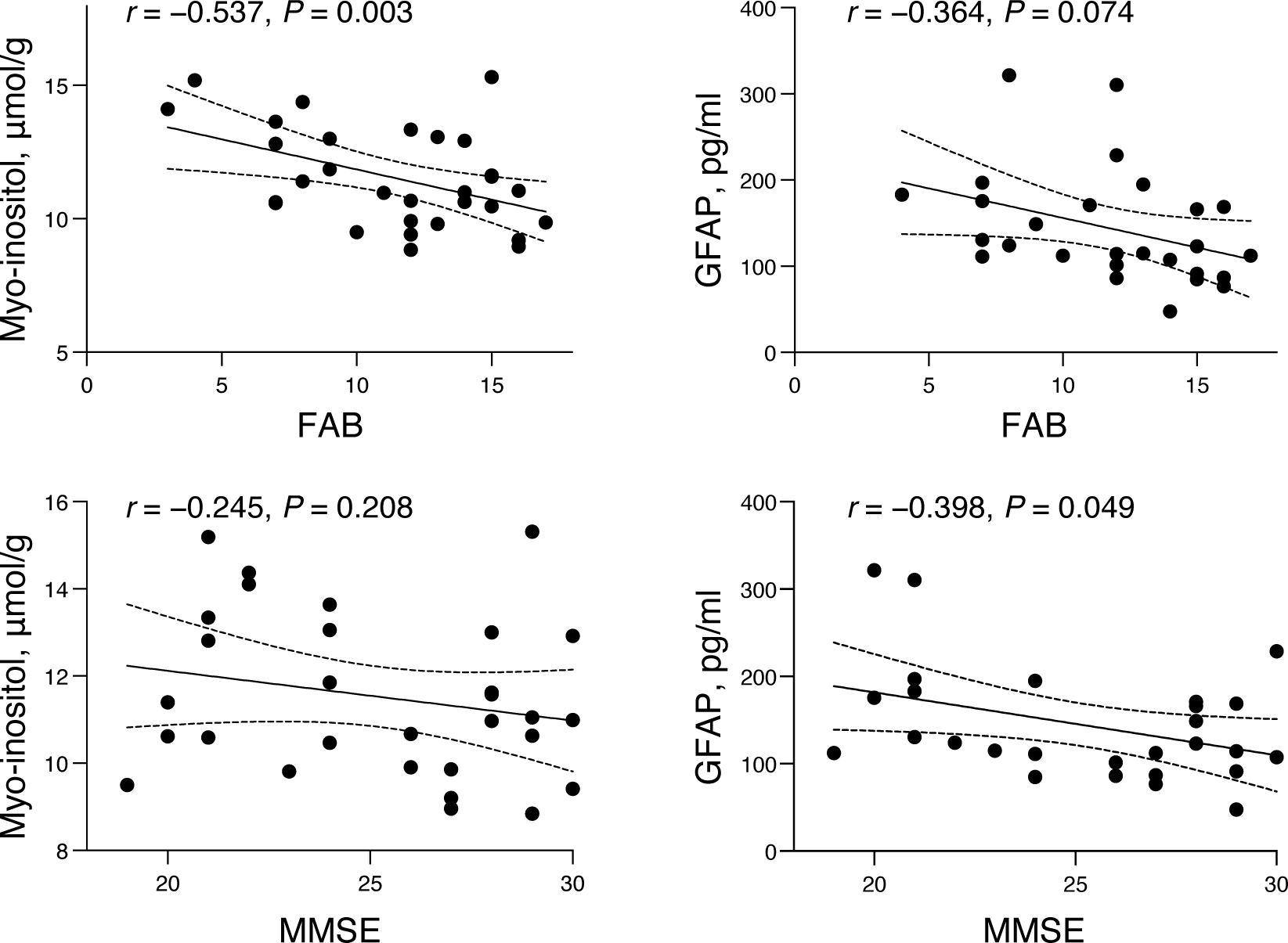
Associations between astrocyte biomarkers and cognitive function scores in progressive supranuclear palsy. The associations between astrocyte biomarkers (myo-inositol in the anterior cingulate cortex and plasma GFAP) and FAB and MMSE scores were assessed using partial correlation analysis, adjusting for age and sex. A linear regression line with a 95% confidence interval was derived from the unadjusted analysis. FAB = Frontal Assessment Battery; GFAP = glial fibrillary acidic protein; MMSE = Mini-Mental State Examination.

### Tau-PET and histological assessment in the anterior cingulate cortex

Finally, we investigated the influence of tau deposition on astrocyte reactivity. First, we examined the ACC for atrophy in PSP; although the VBM analysis revealed atrophy in the midbrain, cerebellum, thalamus, and globus pallidus of participants with PSP, no atrophy was evident in the ACC (Fig. 5A). Accordingly, partial volume correction was not applied for the following PET analysis. Tau-PET retention was assessed in the same ACC VOI as the MRS evaluation (Fig. 5B), revealing no difference in tau-PET retention in the ACC between the PSP group and the HC group (*t* = 1.057, *P* = 0.295) (Fig. 5C). Next, we neuropathologically analysed the ACC region in post-mortem brain samples from another three patients with PSP with Richardson’s syndrome with a disease duration equivalent to that of the current study and healthy controls from different studies. Despite mild tau pathology, manifested by several tufted astrocytes, neurofibrillary tangles, and oligodendroglial coiled bodies, GFAP immunoreactivity was higher in two patients with PSP (PSP-1 and PSP-2) than in HCs (Fig. 6). In the remaining PSP case (PSP-3), GFAP immunoreactivity was almost comparable to that in HCs. The immunoreactivity for synaptophysin was uniformly strong and homogeneous in all cases of PSP and HCs. These results suggest that, in PSP, astrocytes are reactivated in the ACC before the progression of regional tau pathology and neurodegenerative processes.

**Figure 5.**
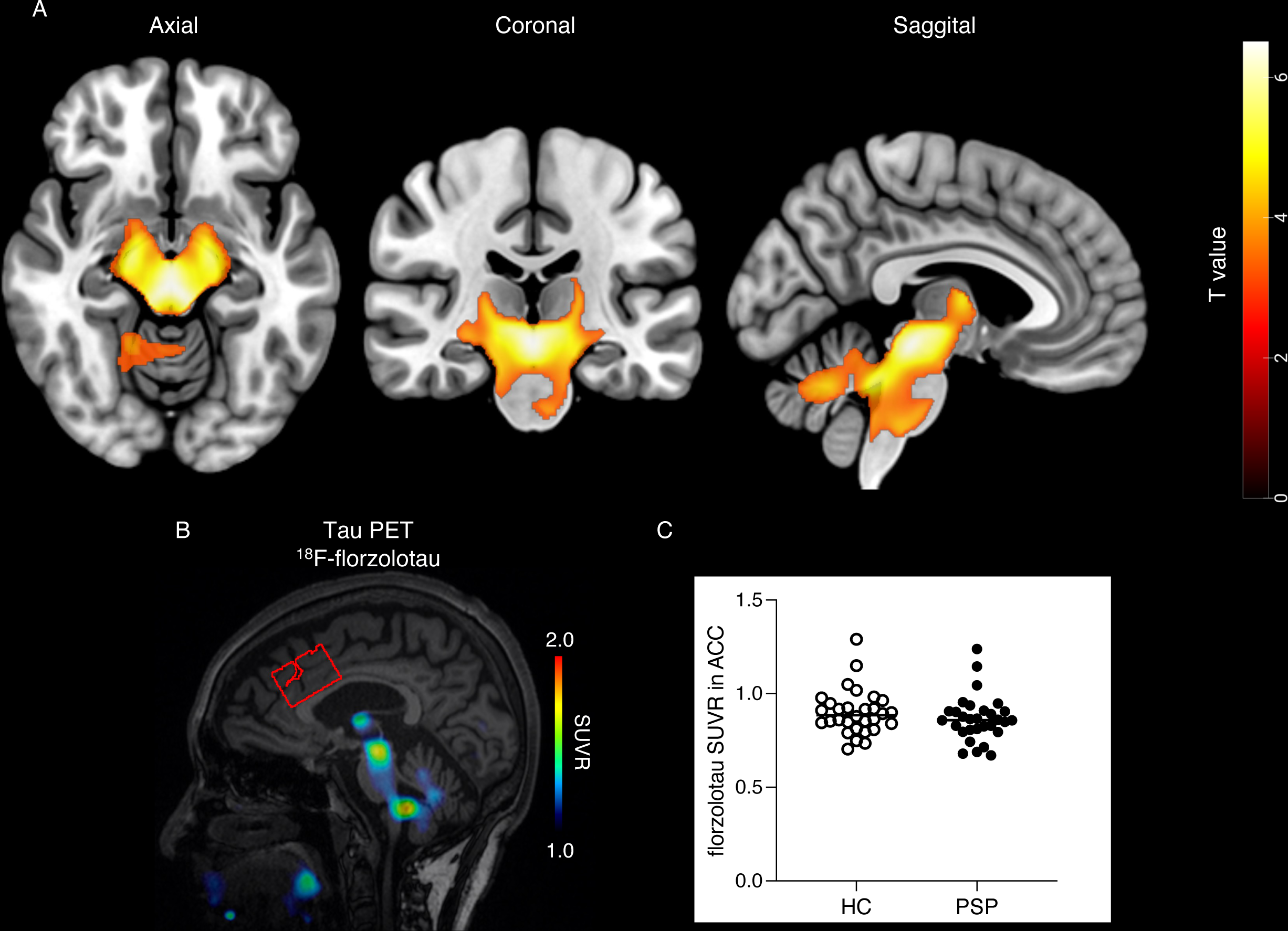
Voxel-based analysis of brain atrophy and tau-PET assessment in the anterior cingulate cortex. **(A)** Voxel-based morphometry analysis showing regions of decreased brain volume relative to healthy control subjects in participants with PSP. The identified regions are displayed on a Montreal Neurological Institute template brain with significance threshold set at *P* < 0.001 at the following Montreal Neurological Institute coordinates: x = −5, y = −22, and z = −13. **(B)** A representative ^18^F-florzolotau-PET image from a participant with PSP with the anterior cingulate cortex voxel utilised to quantify the tau burden. (**C**) Scatter plots of tau-PET retention in the anterior cingulate cortex. Bars indicate median values. ACC = anterior cingulate cortex; HC = healthy control; PSP = progressive supranuclear palsy; SUVR = standardised uptake value ratio.

**Figure 6.**
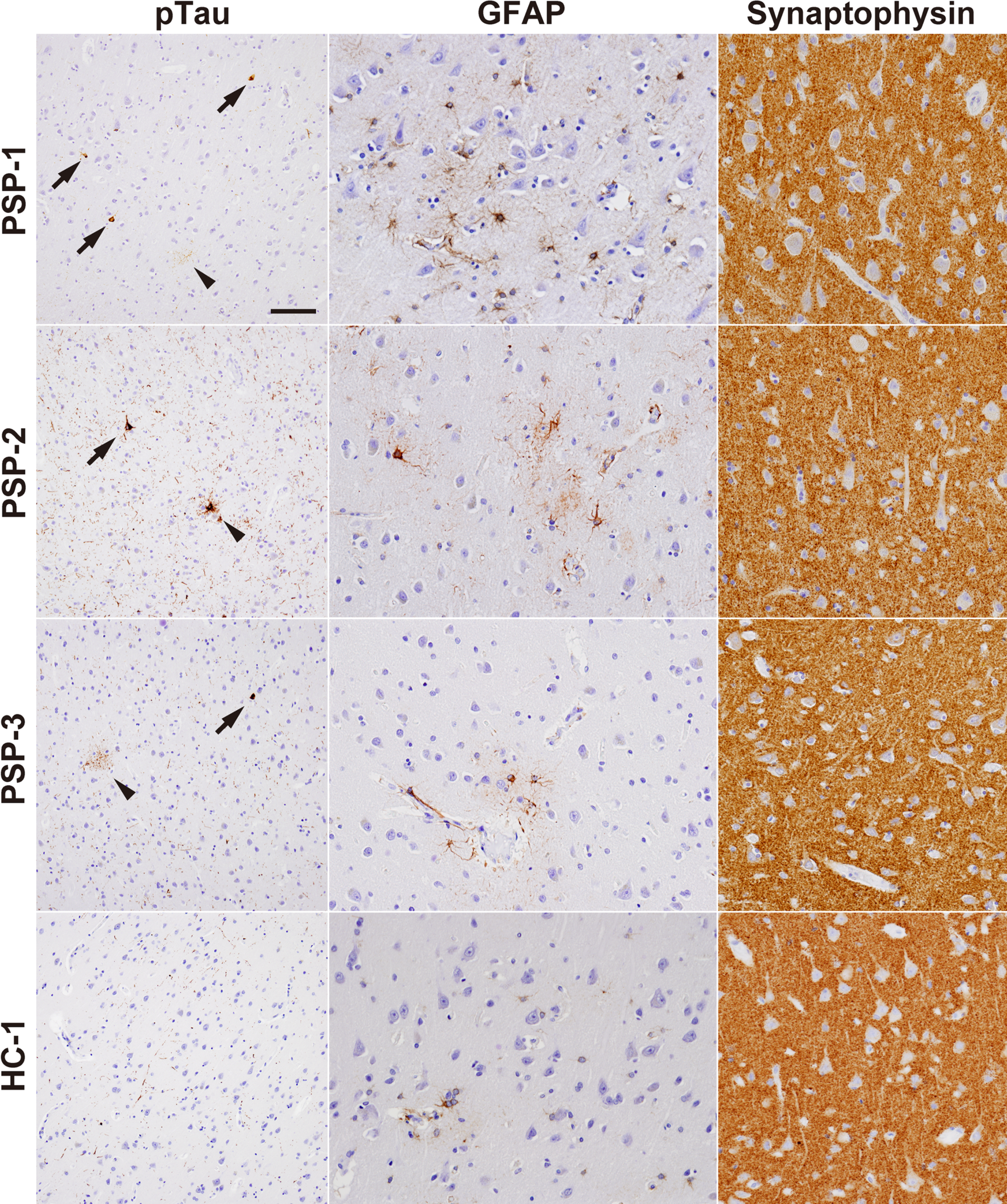
Histological assessment in the anterior cingulate cortex. Neuropathological analysis of the anterior cingulate cortex in autopsy cases of another three patients with PSP with Richardson’s syndrome and healthy controls from different studies. pTau immunohistochemistry showed a small number of tufted astrocytes (arrowheads) and neurofibrillary tangles (arrows) in PSP, but not in controls. In PSP, pTau-positive neuropil threads and oligodendroglial coiled bodies were also observed. pTau-positive neurites were observed as age-related changes in controls. There was an increase in the number and volume of GFAP-positive astrocytes in two PSP cases (PSP-1 and PSP-2) compared to the controls. In the remaining PSP case (PSP-3), GFAP immunoreactivity was comparable to that of controls. Synaptophysin staining demonstrated diffuse immunoreactivity in both patients and controls. Bar: 25 μm for p-tau, 50 μm for GFAP and synaptophysin. HC = healthy control; GFAP = glial fibrillary acidic protein; PSP = progressive supranuclear palsy; pTau = phosphorylated tau.

## Discussion

In this study, we evaluated astrocyte reactivity in patients with PSP with Richardson’s syndrome *in vivo* using myo-inositol in the ACC and plasma GFAP. These astrocyte biomarkers were higher in the PSP group than in the HC group. High myo-inositol and plasma GFAP levels were correlated with cognitive decline in participants with PSP, suggesting that astrocyte reactivity may influence cognitive function. Additionally, increased myo-inositol levels were associated with increased lactate levels in the ACC, an area known to undergo glucose metabolism changes in PSP. Furthermore, with tau-PET and histological evaluations, we discovered that, in the ACC, astrocyte reactivity was observed even with subtle tau deposition.

Elevated myo-inositol levels in the ACC and increased plasma GFAP levels were found in the PSP group compared to the HC group, suggesting astrocyte reactivity. Our results are in line with previous post-mortem pathological studies in PSP.^5,6^ The histopathology of PSP involves widespread gliosis and neuronal loss associated with hyperphosphorylated-tau deposition in astrocytes and neurons mainly in subcortical regions, such as the substantia nigra, subthalamic nucleus, and globus pallidus. Cortical regions, particularly the frontal areas, are also affected from the early stages of the disease and are characterised by predominant astroglial tau pathology.^44^ Although myo-inositol is not specific to astrocytes, as evidenced by the expression of inositol-3-phosphate synthase in various brain cell types (Human Protein Atlas, www.proteinatlas.org),^45^ elevated myo-inositol levels have been linked to reactive astrogliosis in neurodegenerative disorders.^10,12–14^ A recent study in which whole-brain MRS was used revealed a significant increase in myo-inositol levels in the right frontal lobe in patients with PSP, consistent with the present MRS results.^18^ Although only a few studies on blood GFAP concentrations in PSP have been conducted, their results have been inconsistent.^8,9,46^ This inconsistency may be owing to the previous histological observation that tau-positive astrocytes in PSP were minimally immunolabelled with GFAP.^47^ It may also be because GFAP expression tends to increase in response to amyloid; however, this is less likely to be the case in PSP, which is not typically characterised by amyloid accumulation.^48^ Conversely, in this study, plasma NfL levels were markedly elevated in PSP and showed the highest discrimination accuracy between PSP and HCs, consistent with previous studies of plasma biomarkers in PSP.^8,9^ The correlation between the plasma NfL concentration and myo-inositol level suggests a possible involvement of astrocytes in the elevation of plasma NfL. A possible mechanism for this elevation is a contribution of reactive astrocytes to disruption of the blood-brain barrier.^49^

Previous FDG-PET and brain-perfusion SPECT studies have demonstrated the involvement of ACC in patients with PSP with a disease duration comparable to that in our study.^21,50^ Their results support this study in which metabolite levels changed in the ACC in the PSP group. The ACC, situated on the medial surface of the frontal lobes, plays a pivotal role in cognitive and emotional processes, including attention, error detection, and emotion regulation, which are key components of executive function.^22,51,52^ Patients with PSP often experience cognitive and behavioural changes, with frontal lobe dysfunction, including executive function, being a primary cognitive impairment.^23^ A prior rodent study in which chemogenetics and resting-state functional MRI were used revealed that suppressing ACC neuronal activity affects the DMN and behaviour in rats.^53^ These findings suggest that dysfunction of the ACC may underlie the frontal lobe dysfunction observed in PSP. A previous study in which FDG- and ^15^O-PET were combined demonstrated a correlation between frontal hypometabolism and the frontal neuropsychological score in PSP.^54^ Our results, which highlight the relationship between myo-inositol levels in the ACC and the FAB score, which assesses frontal functional deficits, raise the possibility that alterations in astrocyte activity in the ACC may contribute to impaired ACC activity and modulate cognitive function in PSP. This notion aligns with a recent study in which restoration of astrocyte calcium signalling in the ACC normalised cingulate connectivity and resolved behavioural deficits in a mouse model of AD.^55^ Furthermore, the current MRS and histological study of the ACC region in patients with PSP revealed no apparent tNAA decrease or synaptic loss. Our study suggests that functional changes in the ACC without apparent neurodegeneration may influence brain function, an important aspect to consider in the development of therapeutic interventions.

Although aberrant changes in astrocytes can lead to cognitive and behavioural impairments,^56,57^ the underlying mechanisms remain elusive. Given the role of astrocytes in energy metabolism, this study was focused on energy metabolism in the brain. We found that lactate levels were elevated in the ACC, an area known to have altered energy metabolism in patients with PSP.^19,20^ Although research on lactate levels in PSP is limited, one study revealed elevated lactate levels in the CSF of such patients compared to those in HCs.^58^ Additionally, an MRS study revealed lactate peaks in 35% of patients with PSP, which were not present in healthy individuals.^17^ Elevated lactate levels have also been discovered in the brain and CSF of patients with AD and other neuropsychiatric disorders.^11,59,60^ Furthermore, a study involving various mouse models of neuropsychiatric disorders demonstrated a correlation between elevated brain lactate levels and cognitive decline.^61^ These findings indicate that lactate may play a role in the pathophysiology of various neuropsychiatric disorders. Accumulation of lactate can adversely impact brain energy homeostasis. Additionally, high lactate levels can cause a decrease in pH, affecting various cellular processes and functions.^62^ Multiple mechanisms may underlie an elevation in lactate levels. Mitochondrial dysfunction has been implicated in PSP,^63^ suggesting that elevated lactate levels may be due to increased anaerobic metabolism. Moreover, the relationship between myo-inositol and lactate in our study indicates that reactive astrocytes may drive changes in energy metabolism. In a previous *in vitro* study, reactive astrocytes increased calcium signalling, leading to increased glucose uptake and glycolysis, which in turn resulted in elevated lactate levels.^64^ This suggests that abnormal calcium signalling in astrocytes may lead to lactate overproduction. In addition, astrocytes, which are believed to produce a significant amount of lactate in the brain, may transfer it to neurons as an energy source through monocarboxylate transporters (MCTs). This concept is sometimes referred to as the astrocyte-neuron lactate shuttle (ANLS) model.^65^ Several rodent models of AD have demonstrated an altered expression of MCTs, suggesting that dysfunction of the ANLS may contribute to a disruption in lactate utilization and accumulation.^66,67^ However, the methodology used in this study does not allow a detailed discussion of the transfer of lactate from astrocytes to neurons. Nevertheless, tau accumulation in astrocytes can induce changes in astrocyte function, which may lead to high lactate levels in PSP.^68^ Additionally, other cells are also involved in the production and consumption of lactate.^69^ Further studies are needed to elucidate the mechanisms of lactate elevation in PSP. Moreover, although we have focused primarily on brain energy metabolism, astrocytic dysfunction may also play a role in other processes, such as neuroinflammation and synaptic and blood-brain barrier dysfunction, which can result in cognitive deficits.^4,49,57^ Therefore, more research is warranted to uncover in what way the dysfunction of astrocytes is associated with the disease.

Intriguingly, we observed astrocyte reactivity even in the absence of pronounced tau deposition or atrophy. Considering previous pathological studies and VBM analyses, patients with PSP at this stage of the disease in the present study not exhibiting substantial tau accumulation or atrophy in the ACC seems a plausible result.^44,70^ Several factors might have been involved in causing astrocytes to be reactive in this study. One possibility is the influence of even small amounts of tau accumulation as observed in the current histological examination. Aging has also been proposed as a contributing factor for astrocyte reactivity.^71^ Additionally, the observed astrocyte reactivity might have resulted from structural and functional disconnection of the ACC from other distant brain regions.^22^ Future studies using network-based analyses may provide deeper insights into the underlying mechanisms.

In conclusion, in this study, we assessed astrocyte reactivity *in vivo* and provided evidence for the involvement of astrocytes in the pathogenesis of PSP. Myo-inositol, an MRS metabolite, and plasma GFAP were used to reveal astrocyte reactivity, and increased myo-inositol levels in the ACC were associated with frontal lobe dysfunction and altered brain energy metabolism. Our results suggest that, in the ACC, astrocyte reactivity precedes pronounced tau deposition and neurodegenerative processes and modulates brain function in PSP.

## Data availability

The data that support the findings of this study are available from the corresponding author upon reasonable request.

## Acknowledgements

The authors thank all patients and their caregivers for their participation in this study, and clinical research coordinators, PET and MRI operators, radiochemists, and research ethics advisers at QST for their assistance with the current projects. We thank APRINOIA Therapeutics for kindly sharing the precursor of ^18^F-florzolotau. The authors acknowledge the support of Dr Shigeki Hirano and Dr Yoshikazu Nakano at the Chiba University, Dr Shinji Saiki, Dr Taku Hatano, Dr Kenya Nishioka, and Dr Yuta Ishiguro at the Juntendo University, Dr Yoichiro Nishida and Dr Yohsuke Yagi at the Tokyo Medical and Dental Univerisity, Dr Takahiro Takeda at the National Hospital Organization Chiba-Higashi Hospital, and Dr Ikuko Aiba at the National Hospital Organization Higashinagoya National Hospital for patient recruitment.

